# First-Line Systemic Treatment Strategies for unresectable hepatocellular carcinoma : a cost-effectiveness analysis

**DOI:** 10.1101/2022.12.15.22283549

**Authors:** Liting wang, Ye Peng, Shuxia Qin, Xiaomin Wan, Xiaohui Zeng, Sini Li, Qiao Liu, Chongqing Tan

## Abstract

**Background:** Oral multikinase inhibitors and immune checkpoint inhibitors (ICIs) are effective in advanced hepatocellular carcinoma (aHCC) but may increase cost. This study compared the cost-effectiveness of oral multikinase inhibitors and ICIs in first-line patients with aHCC.

**Methods:** A three-state Markov model is established to study the cost-effectiveness of drug treatment from the perspective of Chinese payers. The key outcomes were total cost, quality-adjusted life years (QALYs), and the incremental cost-effectiveness ratio (ICER) in this study.

**Results:** The total costs and QALYs of sorafenib, sunitinib, donafenib, lenvatinib, sorafenib plus erlotinib, linifanib, brivanib, sintilimab plus IBI305, and atezolizumab plus bevacizumab were $9070 and 0.25, $9362 and 0.78, $33814 and 0.45, $49120 and 0.83, $63064 and 0.81, $74814 and 0.82, $81995 and 0.82, $74083 and 0.85, $104188 and 0.84, the drug regimen with the lowest ICER was sunitinib ($551 per QALY), followed by lenvatinib($68869 per QALY). For oral multikinase inhibitors, ICER of levatinib, sorafenib plus erlotinib, linifanib and brivanib compared with sunitinib was $779576, $1534347, $1768971, $1963064, respectively. For ICIs, sintilimab plus IBI305 is more cost effective than atezolizumab plus bevacizumab. The model was most sensitive to the price of sorafenib, the utility of PD, and the price of second-line drugs.

**Conclusion:** For oral multikinase inhibitors, the order of possible treatment options is: sunitinib > lenvatinib > sorafenib plus erlotinib > linifanib > brivanib > donafinb. For ICIs, the order of possible treatment options is: sintilimab plus IBI305 > atezolizumab plus bevacizumab.

## 1. Introduction

Liver cancer is the sixth most common malignancy and the third cause of cancer mortality worldwide[1], Hepatocellular carcinoma (HCC) represents 75%-80% of all liver cancers[1], and the vast majority of HCC is caused by hepatitis B virus (HBV) infection. China accounts for about half of the world’s new HBV infections[2]. Most HCC cases are diagnosed at an advanced stage with a 5-year survival less than 10%[3]. In addition, local resection is not a good option for patients with advanced liver cancer resulting in and very few patients eligible for transplantation[4].

Sorafenib is the first oral multitarget tyrosine kinase inhibitor (TKI) that approved by China in 2007 for the systematic treatment of unresectable advanced HCC (aHCC)[5]. The approval of sorafenib was a response to the positive results from two pivotal Phase III trials called the SHARP and NCT00492752 trials[6, 7], which opened the era of systemic pharmacotherapy for unresectable aHCC[8]. Subsequently, many drugs were successively approved in the first-line setting of unresectable advanced HCC treatment, including oral multikinase inhibitors sunitinib[9], brivanib[10], erlotinib[11], linifanib[12], lenvatinib[13] and donafenib[14], as well as immune checkpoint inhibitors (ICIs) atezolizumab[15] and sintilimab[16]. With the emergence of these novel drugs, physicians and patients face difficulties in determining which is preferable. Recently, a Network Meta-Analysis compared the efficacy and safety of these approved first-line systemic treatment strategies (including the drugs mentioned above)[17]. However, the generalizability of their findings to clinical practice may be limited, in which treatment decision-making need to juggle both cost and efficacy. Therefore, there is a need to conduct a cost-effectiveness analysis to decide the priority of first-line systemic treatment strategies, especially in resource-poor countries like china[18]. Moreover, we use Markov model to build a cost-effectiveness model from the Chinese healthcare system to rank first-line treatments for aHCC.

## 2. Materials and Methods

### 2.1 Analytical Overview

A hypothetical cohort of patients with unresectable aHCC patients who had not previously received systemic therapy was assumed in the model, which mirrored the participants recruited in the SHARP and NCT00492752 trials[6, 7]. Treatment strategies considered in this analysis included the following nine first-line systematic treatments: sunitinib, donafenib, lenvatinib, sorafenib plus erlotinib, sintilimab plus IBI305, linifanib, brivanib, atezolizumab plus bevacizumab, and sorafenib. A three-state Markov model consisting of progression-free survival (PFS), progressed disease (PD) and death was constructed to compare the cost-effectiveness of other eight drugs versus sorafenib in unresectable aHCC (Fig 1). We used a 21-day cycle length and a 10-year time horizon to output the total cost, life-years (LYs), quality-adjusted life years (QALY) associated with each treatment strategies. The cost-effectiveness was measured by using the incremental cost-benefit ratio (ICER), with an ICER lower than the willingness-to-pay (WTP) of (us $37654.50 (defined as the 3 times China’s GDP per capita in 2021)[19]. All costs were expressed in 2022 US dollars using the exchange rate of 1 US dollar equivalent to 6.31 Chinese yuan (March 2022). A 5% annual discount rate was used for both costs and effectiveness[20]. The model was established and analysed by TreeAge Pro (TreeAge Software, Williamstown, MA) and R software version 3.6.1 (https://www.r-project.org/).

**Figure 1:**
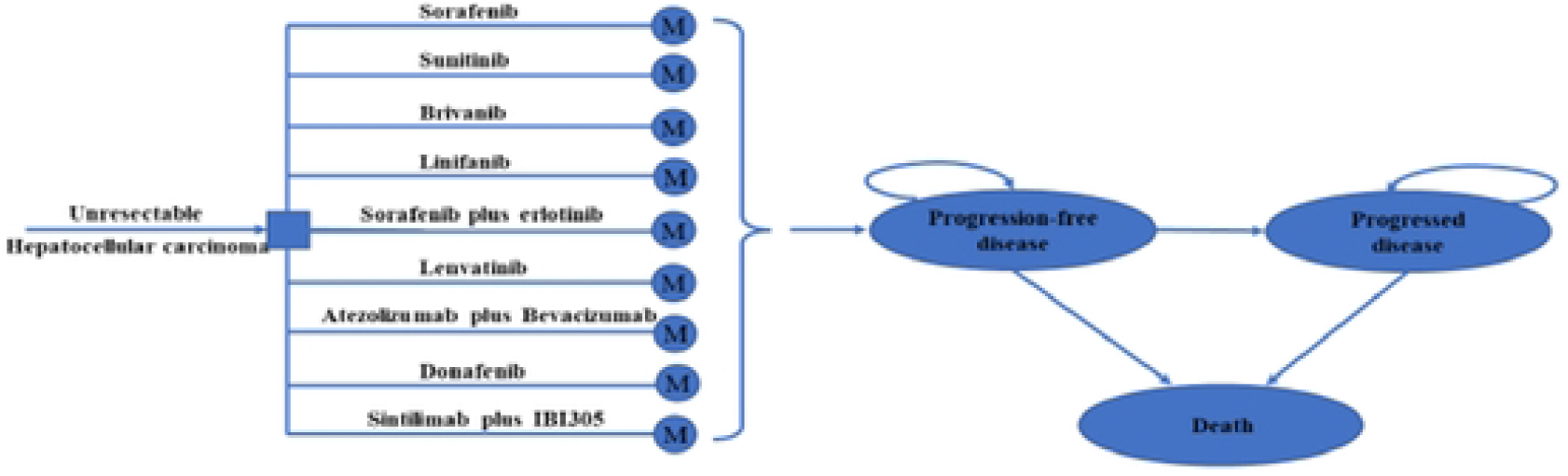
Model structure of a Markov model combining the decision tree. M, Markov.

### 2.2 Clinical Data Inputs

The probability of PFS status transition is estimated from the PFS curve reported in the SHARP and NCT00492752 trials[6, 7], the transition probability from the PFS state to the PD state was calculated using the difference between the OS and PFS curves of the clinical trial, the final probability of death was obtained by subtracting the area under the OS curve of the clinical trial from PFS and OS data of sorafenib patients were derived from SHARP and NCT00492752 trial results[6, 7]. Data points were extracted from OS curves and PFS curves reported in the SHARP and NCT00492752 trial using GetData Graphic Digitizer, version 2.26, and then the algorithm proposed by Guyot et al is used to generate time ranges outside the model[21], finally, Weibull, Exponential, Log-logistic, Log-normal, and Gompertz survival function were fitted, the last survival function selected according to Akaike information criterion (AIC) is Log-Logistic. The survival function for other drugs were adjusted using HR from a network meta-analysis[17]. The subsequent therapy strategies after disease progression were based on guideline recommendations[22]. Key clinical inputs are shown in Table 1.

**Table1:**
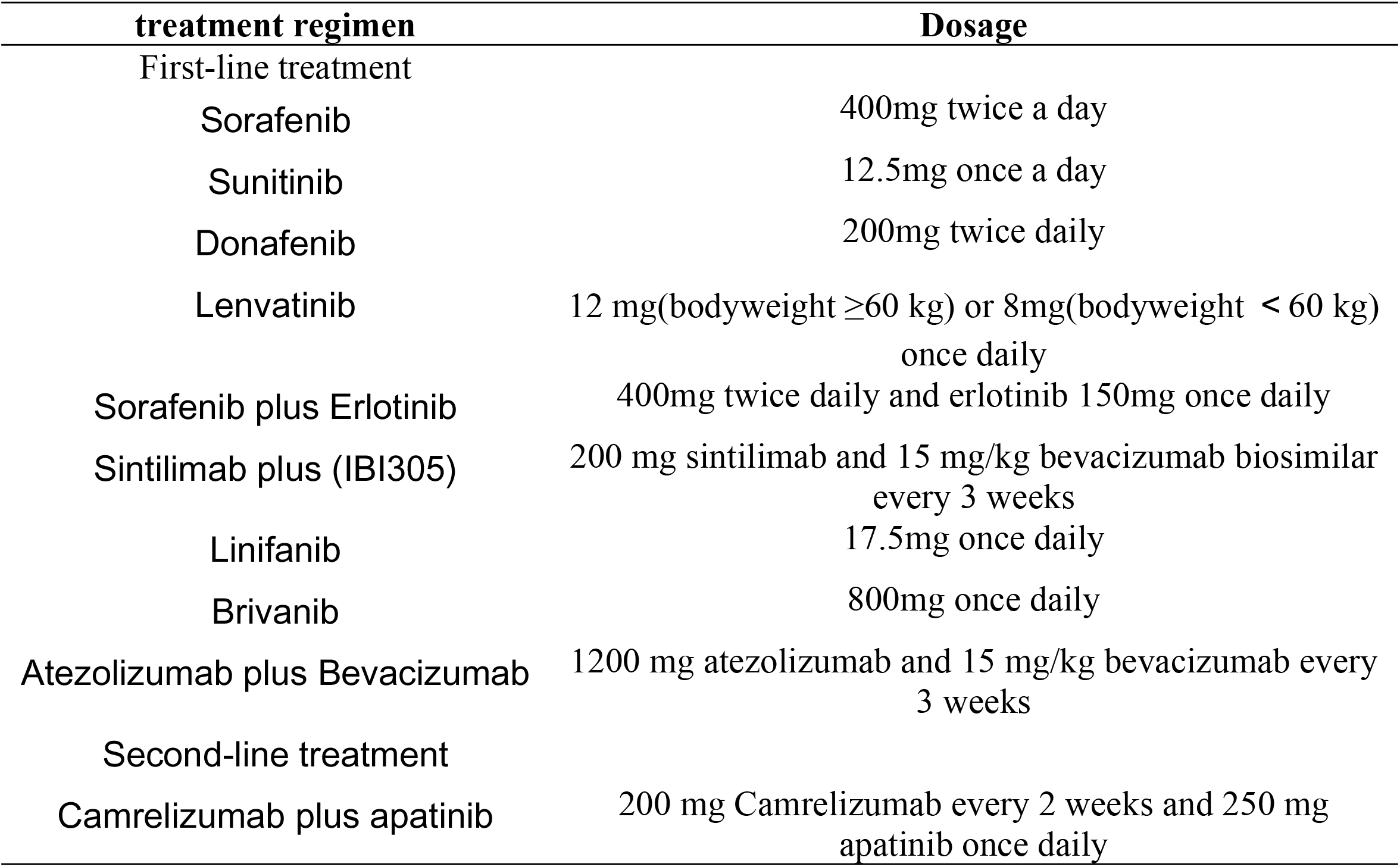
Dosage of treatment regimen

### 2.3 Cost and Utility Inputs

Only direct healthcare costs were covered in this analysis, including drug costs, testing costs and AE costs (table 1). All costs were reported in 2022 US dollars and adjusted to 2022 values based on the consumer Price Index[23].

The study compared nine treatment strategies. The dosage of medication regimen is shown in Table 1, To calculate the dosage of lenvatinib, bevacizumab and IBI305, we modeled the baseline patients as having weighing 60 kg[2]. Patients in all groups were assumed to continued first-line treatment until the disease progressed or unacceptable toxicity. After disease progression, patients were allowed to receive camrelizumab plus apatinib as second-line treatment according to Chinese guideline[23], based on a second-line treatment trial for unresectable hepatocellular carcinoma[24], patients received intravenous camrelizumab 200 mg every 2 weeks plus oral apatinib 250 mg daily. All the information is shown in Table 1

QALYs was measured as a weighted aggregate of health utilities over time. In this analysis, the utility scores assigned to the PFS and PD health state were 0.760 and 0.680[25], respectively. Moreover, experiencing grade 3 or 4 adverse events during each first-line systematic treatment was considered a decrement in health utilities[26], this information is shown in Table1.

### 2.4 Base-Case Analysis

Total expected costs, LYs, QALYs and ICERs were estimated for each first-line systematic treatment. To identify the sensitive parameters that affect our model economic outcomes, we performed both 1-way sensitivity analyses and probabilistic sensitivity analysis. During 1-way sensitivity analyses each parameter were tested within the range of plus or minus 20% of the baseline value, or within the plausible ranges from published literature (S1 table). During probabilistic sensitivity analysis, 10,000 iterations of Monte Carlo simulations were performed by setting appropriate distributions for each parameter. We selected beta distributions for probability, proportion, and preference value parameters, log-normal distributions for the HRs, and gamma distributions for the cost parameters. The cost-effective acceptability curve indicates the possibility of being considered cost-effective at different levels of WTP.

## 3. Results

### 3.1 Base-Case Analysis

Table 2 summarizes the base-case results. the atezolizumab plus bevacizumab group had the highest total cost (Us $104,188), the sorafenib group had the least total cost (US $9362); The QALYs obtained by atezolizumab plus bevacizumab, sintilimab plus IBI305, brivanib, linifanib, sorafenib plus erlotinib, lenvatinib, donafenib and sunitinib relative to sorafenib treatment group were respectively 0.84, 0.85, 0.82, 0.82, 0.81, 0.83, 0.45 and 0.78, respectively, increased by 0.59, 0.56, 0.57, 0.59, 0.57, 0.58, 0.20 and 0.53 QALYs. When patients received oral multikinase inhibitors as first-line systematic treatments, compared with sorafenib, the ICER for lenvatinib, sorafenib plus erlotinib, linifanib, brivanib versus sunitinib were $779576, $1534347, $1768971and $1963064 per QALY, respectively. When patients received immune checkpoint inhibitors (ICIs) as first-line systematic treatment, atezolizumab plus bevacizumab was dominated by sintilimab plus bevacizumab IBI305. Detailed data are shown in Table 3.

**Table2:** Base case results.

| Drug Name | Lys | Total cost(\$) | QALYs | incrC | incrE | ICER (per QALY) |
| --- | --- | --- | --- | --- | --- | --- |
| Sorafenib | 0.336 | 9070.170 | 0.249 | / | / | / |
| Sunitinib | 1.029 | 9362.394 | 0.779 | 292.224 | 0.530 | 551.305 |
| Donafenib | 0.630 | 33814.382 | 0.453 | 24744.213 | 0.204 | 121059.073 |
| Lenvatinib | 1.176 | 49120.781 | 0.830 | 40050.612 | 0.582 | 68869.402 |
| Sorafenib Plus Erlotinib | 1.176 | 63064.570 | 0.814 | 53994.400 | 0.565 | 95545.057 |
| Sintilimab plus (IBI305) | 1.192 | 74083.924 | 0.845 | 65013.754 | 0.597 | 108970.232 |
| Linifanib | 1.169 | 74814.336 | 0.816 | 65744.166 | 0.568 | 115760.370 |
| Brivanib | 1.168 | 81995.796 | 0.816 | 72925.627 | 0.567 | 128527.131 |
| Atezolizumab plus Bevacizumab | 1.192 | 104188.761 | 0.843 | 95118.591 | 0.594 | 160049.163 |

Lys: life-years; QALYs: quality-adjusted LYs; ICER: incremental cost-effectiveness ratio.

**Table3:** Base case results.

| Treatment strategies | Cost | incrC | QALY | incrE | Scheme comparison | ICER |
| --- | --- | --- | --- | --- | --- | --- |
| Sorafenib | 9070 | / | 0.25 | / | / |  |
| Sunitinib | 9362 | 292 | 0.78 | 0.53 | Sorafenib | 551 |
| Donafenib | 33814 | 24744 | 0.45 | 0.20 | Sorafenib | 121059 |
| Lenvatinib | 49120 | 40050 | 0.83 | 0.58 | Sorafenib | 68869 |
| Sorafenib Plus Erlotinib | 63064 | 53994 | 0.81 | 0.57 | Sorafenib | 95545 |
| Linifanib | 74814 | 65744 | 0.82 | 0.59 | Sorafenib | 115760 |
| Brivanib | 81995 | 72925 | 0.82 | 0.57 | Sorafenib | 128527 |
| Sintilimab plus (IBI305) | 74083 | 65013 | 0.85 | 0.56 | Sorafenib | 115760 |
| Atezolizumab plus Bevacizumab | 104188 | 95118 | 0.84 | 0.59 | Sorafenib | 160049 |

incrC: incremental cost, QALYs: quality-adjusted-life-years ; incrE: incremental effect; ICER: incremental cost-effectiveness ratio.

**Table4:** Results of oral multikinase inhibitor group and ICI group

| Drug Name | Total cost(\$) | QALYs | ICER (per QALY) |
| --- | --- | --- | --- |
| oral multikinase inhibitors |  |  |  |
| Sunitinib | 9362 | 0.78 | / |
| Lenvatinib | 49120 | 0.83 | 7795762 |
| Sorafenib Plus Erlotinib | 63064 | 0.81 | 1534347 |
| Linifanib | 74814 | 0.82 | 1768971 |
| Brivanib | 81995 | 0.82 | 1963064 |
| Donafenib | 33814 | 0.45 | dominated |
| immune checkpoint inhibitors (ICI) |  |  |  |
| Sintilimab plus IBI305 | 74083 | 0.85 | / |
| Atezolizumab plus Bevacizumab | 104188 | 0.84 | dominated |

QALYs, quality-adjusted life-years. ICER, incremental cost-effectiveness ratio.

### 3.2 Sensitivity Analysis

Results of the one-way sensitivity analysis showed that for sunitinib versus sorafenib, the ICER was particularly sensitive to the price of sorafenib; for donafenib, sorafenib plus erlotinib, linifanib, brivanib, sintilimab plus IBI305 and atezolizumab plus bevacizumab versus sorafenib, the utility of PD was the most sensitive parameter; for lenvatinib versus sorafenib, the price of second-line drugs was the most sensitive parameter, for sunitinib, sorafenib plus erlotinib and brivanib versus lenvatinib, the utility of PD was the most sensitive parameter, for donafenib versus lenvatinib, the price of second-line drugs was the most sensitive parameter. Other sensitive parameters are shown in S1 Fig. With the increase of the WTP thresholds, the possibility of cost-effectiveness among different treatment drugs increased, but the cost-effectiveness of all drug treatment regimens except sorafenib was less than 50%, as shown in Fig 2.

**Figure 2:**
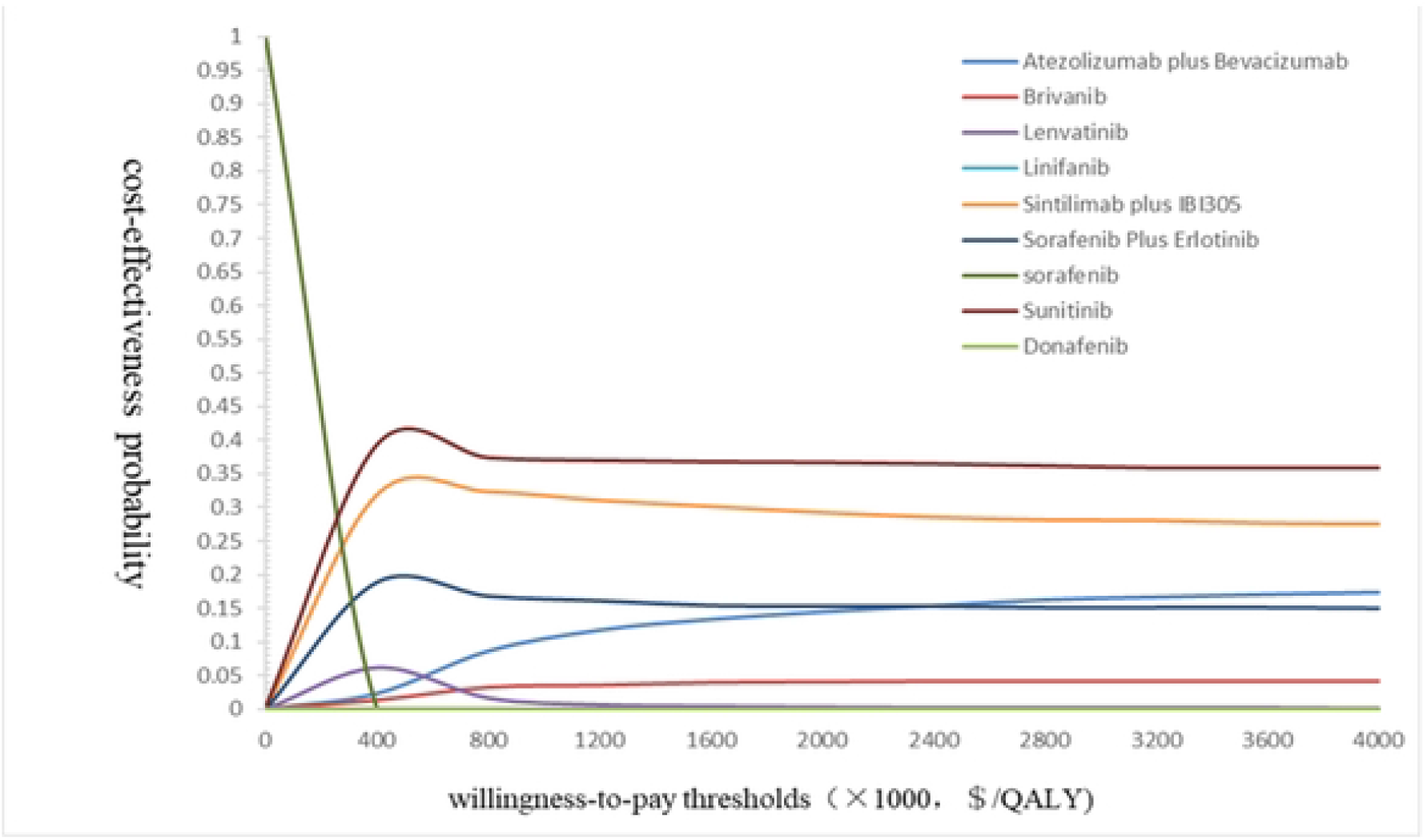
Cost effectiveness acceptability curves. QALYs, quality-adjusted life-years.

## 4. Discussion

We used the three-state Markov model to analyze the cost-effectiveness of eight first-line treatment plans. The drugs of the eight treatment plans are mainly oral multikinase inhibitors and immune checkpoint inhibitors. Compard with standard sorafenib treatement, all other first-line treatements was superior in improving survival, among them, sunitinib treatment plan is a cost-effectiveness choice, with an ICER of $551/QALYs below the WTP threshold of $37654.50/QALYs. China also began negotiating with pharmaceutical companies over the price of cancer drugs after establishing the National Medical Safety Administration (NHSA) in May 2018, Among them, Sunitinib, Lenvatinib and Sintilimab have entered the NHSA, and we look forward to more drugs. For oral multikinase inhibitors, our secondary study found that the QALY value of lenvatinib was 0.83, which was the largest, and this first-line treatment plan certainly had the best long-term efficacy. Moreover, the indication for lenvatinib to be included in the medical insurance list was advanced hepatocellular carcinoma, which could reduce the economic burden of many patients. We assumed that the patient’s weight was 60kg and the dose was 8mg, and didn’t consider the 12mg dose for the patient’s weight over 60kg, Hongfu Cai et al. studied that both treatment regimens weighing less than 60kg and more than 60kg were cost-effective, and sensitivity analysis showed that weight was not a major influencing factor[27]. So lenvatinib has a greater chance of being more cost-effective than the other four drugs. In the REFLECT clinical trial, levatinib was reported to be more effective than sunitinib, brivanib, linifanib, and erlotinib plus sorafenib[13]. Hongfu Cai et al. proved the cost-effectiveness of lenvatinib versus sorafenib from the perspective of China, Brandon M Meyers et al. demonstrated the cost-effectiveness of lenvatinib versus sorafenib from a Canadian perspective[28]. Christopher Sherrow et al. proposed that lenvatinib is the most cost-effective first-line treatment when only oral therapy is considered in the sequencing of systematic treatment options for aHCC[28], this is consistent with our finding.

When comparing the two ICI schemes, Sintilimab plus IBI305 has a greater chance of being cost-effective relative to atezolizumab plus bevacizumab, Feng Wen et al. found that Atezolizumab plus bevacizumab was not cost-effective from the Perspective of China[25], although the FDA’s recommendation to use atezolizumab plus bevacizumab in patients with aHCC who have not previously received systemic treatment has ushered in a new era of systemic treatment for hepatocellular carcinoma[29], the high cost is an issue that cannot be ignored, especially in developing countries. Ye Peng et al. and Ting Zhou et al. demonstrated the cost-effectiveness of sintilimab plus bevacizumab biosimilar from a Chinese perspective for aHCC patients[4, 30]. Similarly, compared with other ICI, FDA of the United States and National Medical Products Administration (NMPA) of China also confirmed that sintilimab has similar anti-tumor effects, better safety and obvious economic advantages, this is a valuable finding for Chinese patients. Therefore, there are discrepancies between the results of the sintilimab study and those of our article, which may be due to differences in second-line regimens. From our study, we can know that the price of domestic anticancer drugs is much lower than that of imported anticancer drugs, so the cost-effectiveness analysis of them is of great significance for reducing national health expenditure. As can be seen from the above, domestic sintilimab has been included in the National Reimbursement Drug List (NRDL), but the indication is Hodgkin’s lymphoma. With the success of the phase III clinical trial Orient-32, we believe that the indication for unresectable advanced HCC will soon be included in the NRDL.

Notably, as shown in ICER values (S1 table), only sunitinib was cost-effective according to China’s WTP threshold in the range of $38,498.89 /QALY, and no other systematic drug regimen has been deemed cost-effective, the calculated ICER values were all exceed this threshold. The main reason is that the cost may be affected by the second-line treatment, which can be seen from the results of one-way sensitivity analysis, apart from sunitinib, the biggest factors affecting ICER of other treatment drugs are the cost of second-line drugs and the utility of PD. Nassir A. Azimi et al. found that if ICER is greater than an increase of $166,000 /QALY, researchers are generally opposed to implementing this treatment regimen. If ICER is in the range of $61,500 - $166,000 /QALY, cost-effectiveness is ambiguity[31], so researchers come to different conclusions in this range. Our study clearly Outlines the cost of each first-line drug treatment regimen to select the most cost-effective regimen for Chinese patients.

There are some limitations to our study. First, safety and efficacy data used in our study were extracted from published trials, any biases in these trails may inevitably affect our results; Second, the potential heterogeneity across different patient populations was not considered in the model, resulting in no subgroup analysis, we did not consider effective biomarkers, which can predict the outcome of immunotherapy, and are critical in the logical context of optimal cost-effectiveness; Third, the assumptions of the data also limited our analysis, for second-line treatment, due to the lack of real world data validation, we assume that camrelizumab plus apatinib system drug therapy recommended by Chinese hepatocellular carcinoma guidelines is considered, because this article is from the perspective of China, hoping to facilitate Chinese patients, there is no denying that it will cause deviation in clinical practice, but the focus of our study is the first-line drug treatment, the unification of second-line treatment, better see the difference of first-line drug treatment; we also assume that the threshold for sensitivity analysis is 20% above or below, which may affect the reality when interpreting uncertainty, but in similar studies, this is an acceptable range[32]; Finally, our analysis does not address the effects of different payment options, which is a more practical issue for most policymakers and patients, although it is not the focus of the study.

## 5. Conclusions

Our study demonstrates the possibility of cost-effectiveness of different drug regimens and provides more different treatment ideas for advanced HCC patients in China.

## Data Availability

All date files are available from the publish paper

## Conflict of Interest

The authors declare that the research was conducted in the absence of any commercial or financial relationships that could be construed as a potential conflict of interest.

## Author Contributions

Study design and supervision: QL, CT; Data analysis and interpretation: LW, SL, YP; Data collection: XW, XZ, SQ; Manuscript writing: LW; Final approved of manuscript: All authors.

## Funding

1. The work was supported by grants from the National Natural Science Foundation of China (grant numbers: 82073818)
2. The Fundamental Research Funds for the Central Universities of Central South University(grant numbers:2022ZZTS0282)
3. Supported by Hunan Provincial Innovation Foundation For Postgraduate (grant numbers: CX20220364)

## Notes

### Competing Interest Statement

NO authors have competing interests Enter: The authors have declared that no competing interests exist.

### Funding Statement

YES - Specify the role(s) played.

### Author Declarations

No ethical information is required

